# Prevalence of long covid-19 among pediatric age group in Duhok city, Kurdistan region, Iraq

**DOI:** 10.1101/2023.08.05.23293695

**Authors:** Rojeen Chalabi Khalid, Tamara Bassam Jamal, Sara Ardalan Mahdi, Abdullah Saeed Mustafa

## Abstract

**Background:** A range of persistent symptoms that can develop in some people after they have recovered from acute COVID-19, is known as Long COVI-19. It can affect people of all ages and severity of initial illness, including those who had mild or asymptomatic infections.. Dealing with Long COVID-19 can be challenging, and the best course of action will depend on the specific symptoms and individual needs of the patient. This study aims to detect the prevalence of long covid-19 among the children who tested positive for IgG test. If IgG antibodies are detected in a person’s blood sample, it suggests that they have been infected with SARS-CoV-2 at some point in the past and their immune system has responded by producing antibodies against the virus.

**Material and Methodology:** From (October 22^nd^ till December 4^th^ 2022) the data of this study had been collected through a face-to-face interview with withdrawing blood samples for serum Immunoglobin-G test in laboratory of (General zakho Teaching Hospital in Zakho) and (Hevi Pediatric Teaching Hospital in Duhok). A total number of 330 children aged between 5-12 ages participated in this study. Moreover, If IgG antibodies are detected in a person’s blood sample, it suggests that they have been infected with SARS-CoV-2 at some point in the past and their immune system has responded by producing antibodies against the virus.

**Results:** (Fatigue 12/ 85.7%), (cough 10/ 71.4%), (post exertional malaise 5/ 35.7%) were the most detected symptoms among the 14 positive patients. Followed by (headache, dizziness, hair loss, loss of appetite, loss/change in smell and taste, difficulty in sleep, mood change, abdominal pain, change in bowel habits, chest pain) to lesser extent.

**Conclusion:** long-term sequelae of Covid-19 now is becoming a challenge that needs more continued research and collaboration among healthcare providers, researchers, and patients are essential. Long COVID-19 is a public health concern that requires ongoing attention and resources, as well as support for those who are experiencing its debilitating effects. Out of 330 children only 4.6% (14 children) were experiencing long covid-19 symptoms for more than 4 weeks after acute infections in Duhok city.

## 1. Introduction

In recent years, there has been a concerning trend of reemerging viral infections, posing significant challenges to global public health and necessitating a comprehensive understanding and effective strategies to combat these resurgent threats (1-3). The COVID-19 pandemic has had a profound impact on the world, leading to significant social, economic, and political upheaval. COVID-19 is a contagious respiratory illness caused by the SARS-CoV-2 virus (4). The term “COVID-19” stands for “Coronavirus Disease 2019” and was coined by the World Health Organization (WHO) In December 2019: The first cases of a new coronavirus are reported in Wuhan, China. The virus is later named SARS-CoV-2. Symptoms of COVID-19 can range from mild (fever, cough, fatigue) to severe (shortness of breath, pneumonia, acute respiratory distress syndrome), and can lead to hospitalization and even death (5, 6). The World Health Organization (WHO) in January 2020 declares a Public Health Emergency of International Concern (PHEIC) as the number of cases in China rapidly increases. Many countries implement travel restrictions and other measures to contain the spread. By March 2020: The pandemic is declared by the WHO. Governments around the world implement strict lockdowns, social distancing measures, and other public health interventions to slow the spread of the virus. From July-December 2020: Vaccines begin to be authorized and distributed, starting with healthcare workers and high-risk populations. Despite this, the virus continues to spread rapidly, and many countries experience a surge in cases and deaths. The possibility of reinfection worsens the pandemic and increased the risk of prolong crisis (7-10). The pandemic had a deleterious impact on the health system in Iraq and elsewhere (11-16).

The World Health Organization (WHO) reported that the highest number of new cases per week were being reported in the Americas, followed by Europe and Southeast Asia. The incidence of COVID-19 in children has varied over time and across different geographic locations. While children can and do contract COVID-19, they have generally been less likely to experience severe illness or require hospitalization than adults. The main reported risk factors for the pediatric population to be infected with COVID-19 were close contact with a family member with an infection and a history of travel or residence in an endemic area. Until now, no study could identify the prevalence of co-morbidities in children infected with COVID-19. It’s important to note that while children may be less likely to experience severe illness from COVID-19, they can still transmit the virus to others, including those who are at higher risk of developing severe illness Vaccination is an important tool for protecting children and others from COVID-19(17).

The pathophysiology of COVID-19 is complex and not yet fully understood, but it is believed to involve a combination of viral infection, immune response, and inflammation(18-20).

COVID-19 primarily spreads through respiratory droplets that are expelled when an infected person talks, coughs, or sneezes. These droplets can then be inhaled by others who are in close contact with the infected person, or they can land on surfaces and objects, where they can be picked up by others who touch these surfaces and then touch their face. Symptoms of COVID-19 can vary widely from person to person, but may include (Fever or chills, Cough, Shortness of breath or difficulty breathing, Fatigue, Muscle or body aches, Headache, New loss of taste or smell, Sore throat, Congestion or runny nose, Nausea or vomiting, Diarrhea) It’s important to note that not everyone who has COVID-19 will experience symptoms, and some people may only have mild or no symptoms at all. However, even asymptomatic individuals can still transmit the virus to others, however there are several variants of the SARS-CoV-2 virus that causes COVID-19, some of which are considered “variants of concern” or “variants of interest” because they may have an increased ability to spread, cause more severe disease, or potentially evade immunity generated by vaccines or previous infections(15).

Long COVID, also known as post-acute sequelae of SARS-CoV-2 infection (PASC), refers to a range of persistent symptoms that can develop in some people after they have recovered from acute COVID-19. Long COVID can affect people of all ages and severity of initial illness, including those who had mild or asymptomatic infections(21, 22). Long COVID can have a significant impact on a person’s quality of life and ability to carry out daily activities, and it is not yet clear why some people develop these persistent symptoms while others do not. Research is ongoing to better understand the mechanisms of Long COVID and develop effective treatments(4). People who experience post-COVID conditions most commonly report: Fatigue, Shortness of breath, Chest pain or tightness, Joint or muscle pain, Difficulty sleeping, Headache, Brain fog or difficulty concentrating, Depression or anxiety, Loss of sense of smell or taste, Digestive symptoms such as nausea or diarrhea. Risk factors for long covid-19 could be less appreciated, as the risks associated with covid-19 appears not to have a significant association with risk of long covid, however male sex, age, and pre-existing conditions including obesity, diabetes, and cardiovascular disease have shown no association with the risk of developing long covid. However, pre-existence of asthma has been found to be significantly associated with long covid. This study aims to detect the prevalence of long covid-19 among the children who tested positive for IgG test(4). If IgG antibodies are detected in a person’s blood sample, it suggests that they have been infected with SARS-CoV-2 at some point in the past and their immune system has responded by producing antibodies against the virus.

## 2. Material and methodology

### 2.1 Study design

This cross-sectional study was conducted from (October 22^nd^ till December 4^th^ 2022) Among pediatric age group individual of Zakho and Duhok cities, Kurdistan region, Iraq. through a face-to-face interview with blood samples to be withdrawn for serum Immunoglobin-G test in laboratory of (General zakho Teaching Hospital in Zakho) and (Hevi Pediatric Teaching Hospital in Duhok). A total number of 330 children age between 5-12 years old have been recruited in this study. The data of this study was collected through administering a questionnaire. The scheme of questions in which the subject has to go through was divided into 2 main parts, the first part contains demographic data of the child and mother such as (name, age, sex, address, phone number, socioeconomic status, mother occupation, number of family member, birth weight, premature labor, feeding, any known chronic illnesses, current weight, current height, BMI, immunization status, mother education level) the second part contains questions regarding covid-19 infection such as (history of documented covid-19, severity of infection, evidence of pneumonia symptoms, family history of documented covid-19, symptoms of infection during the active state, symptoms of infection after 4 week from infection).

### 2.2 Statistical analysis

The collected data of this study had been analyzed by SPSS software. Chi-square test was used to study the association between the different variables. A p-value of 0.05 or less was considered statistically significant.

### 2.3 Inclusion criteria

This study’s data included all children aged between (5-12) years who tested positive to serum IgG test in Zakho and Duhok cities.

### 2.4 Exclusion criteria

All children aged between (5-12) years who tested negative to serum IgG test and children below (5) years, above (12) years in Zakho and Duhok cities had been excluded.

### 2.5 Ethical approval

This study was approved by the Ethics and scientific committee of college of medicine, university of zakho, Kurdistan region, Iraq. All the subjects included in this study have been informed that their personal data will not be shared, and a formal consent has been taken from each individual.

## 3. Results

### 3.1 Basic Demographic Characteristics

The present study aimed to investigate the phenomenon of long Covid-19, which is a persisting set of symptoms and complications that occur after the acute phase of Covid-19 has resolved. The cases of our study had been taken in laboratory of (General Zakho Teaching Hospital) and (Hevi Pediatric Teaching Hospital), in Zakho and Duhok cities, respectively. the total number of children who participated in this study were 330 children aged between (5-12) years old. Among them 170 (51.5%) were male and 160 (48.5%) were female. 302 participants were positive IgG (male 156 / 51.7%) (female 146/ 48.3%), in other hand only 28 participants were negative IgG (male 14/ 50.0 %) (female 14/ 50.0%). Among those who were positive IgG (14 participants 4.6%) alone showed the symptoms for long covid-19. While the other (288 participants) did not show such symptoms for long covid-19. (***See. Table 1***.)

**Table 1:** Characteristics of long Covid patients.

| Characteristics | Long Covid |  |
| --- | --- | --- |
|  | N | % |
| Age Group |  |  |
| 5-7 | 5 | 35.7 |
| 8-10 | 6 | 42.9 |
| 11-12 | 3 | 21.4 |
| Gender |  |  |
| Male | 8 | 57.1 |
| Female | 6 | 42.9 |
| Birth Weight |  |  |
| less than 2 | 1 | 7.1 |
| 2-3 | 2 | 14.3 |
| 4-5 | 11 | 78.6 |
| Socioeconomic Status |  |  |
| Poor | 4 | 28.6 |
| Accepted | 6 | 42.9 |
| Good | 4 | 28.6 |
| Mother Occupation |  |  |
| Employ | 2 | 14.3 |
| House wife | 12 | 85.7 |
| Premature Labor |  |  |
| Yes | 2 | 14.3 |
| No | 12 | 85.7 |
| Feeding |  |  |
| Brest feeding | 5 | 35,7 |
| Formula feeding | 2 | 14.3 |
| Mixed feeding | 7 | 50 |
| Any evidence of pneumonia |  |  |
| Yes | 3 | 21.4 |
| No | 11 | 78.6 |
| Number of family members |  |  |
| less than 3 | 2 | 14.3 |
| 4 to 6 | 6 | 42.9 |
| 7 to 9 | 4 | 28.6 |
| 10 to 12 | 1 | 7.1 |
| more than 13 | 1 | 7.1 |

### 3.2 long Covid-19 patients associated with different variables

Our data illustrated the association of positive long covid-19 patients with different variable including (Age, Gender, Birth Weight, Socioeconomic Status, Mother Occupation, Premature Labor, Feeding, Any evidence of pneumonia, Number of family members). And it went as follow (age 5-7 years 5 / 35.7%), (age 8-10 years 6 / 42.9%), (age 11+ years 3/ 21.4%), (age P-value 0.775). (Male gender 8 / 57.1%), (Female gender 6/ 42.9%), (gender P-value 0.832). (birth weight of <=1 1 / 7.1%), (birth weight of 2-3 2/ 14.3%), (birth weight of 4-5 11/ 78.6%),(birth weight P-value 0.064). (Socioeconomic Status poor 4/28.6%), (Socioeconomic Status accepted 6/ 42.9%), (Socioeconomic Status good 4/ 28.6%), (Socioeconomic Status P-value 0.904). (

Mother Occupation /employ 2/ 14.3%), (Mother Occupation/ house wife 12/ 85.7%), (mother occupation P-value 0.981). (Premature Labor /yes 2/ 14.3%), (Premature Labor/ no 12/ 85.7%), (premature labor P-value 0.039). (feeding/ Brest feeding 5/35.7%), (feeding/ formula feeding 2/14.7%), (feeding/ mixed 7/50.0 %), (feeding P-value 0.101). (Any evidence of pneumonia / yes 3/21.4%), (Any evidence of pneumonia / no 11/78.6%), (Any evidence of pneumonia P-value 0.477). (Number of family members <=3 2/14.3%), (Number of family members 4-6 6/42.9%), (Number of family members 7-9 4/28.6%), (Number of family members 10-12 1/7.1%), (Number of family members 13+ 1/7.1%), (Number of family members P-value 0.803). (***See. Table 2***)

**Table 2.** Associations of long covid with different variables.

| Characteristics | Positive IgG (N=288) |  | Long Covid Positive IgG (N=14) |  | *P-value* |
| --- | --- | --- | --- | --- | --- |
|  | N | % | N | % |  |
| **Age group** |  |  |  |  |  |
| *5-7* | 126 | 43.8 | 5 | 35.7 | 0.605 |
| *8-10* | 106 | 36.8 | 7 | 50 |  |
| *11+* | 58 | 19.4 | 2 | 14.3 |  |
| **Gender** |  |  |  |  |  |
| *Male* | 147 | 51 | 9 | 64.3 | 0.51 |
| *Female* | 141 | 49 | 5 | 35.7 |  |
| **Birth Weight** |  |  |  |  |  |
| *<=1* | 3 | 1 | 1 | 7.1 | 0.199 |
| *2-3* | 24 | 8.3 | 2 | 14.3 |  |
| *3-4* | 226 | 78.5 | 11 | 78.6 |  |
| *4-5* | 33 | 11.5 | 0 | 0 |  |
| *5+* | 2 | 0.7 | 0 | 0 |  |
| **Socioeconomic Status** |  |  |  |  |  |
| *Poor* | 59 | 20.5 | 5 | 35.7 | 0.278 |
| *Accepted* | 182 | 63.2 | 6 | 42.9 |  |
| *Good* | 47 | 16.3 | 3 | 21.4 |  |
| **Mother Occupation** |  |  |  |  |  |
| *Employ* | 32 | 11.1 | 3 | 21.4 | 0.239 |
| *House wife* | 256 | 88.9 | 11 | 78.6 |  |
| **Premature Labor** |  |  |  |  |  |
| *Yes* | 15 | 5.2 | 2 | 14.3 | 0.15 |
| *No* | 273 | 94.8 | 12 | 85.7 |  |
| **Feeding** |  |  |  |  |  |
| *Brest feeding* | 69 | 24 | 4 | 28.6 | 0.7 |
| *Formula feeding* | 69 | 24 | 2 | 14.3 |  |
| *Mixed feeding* | 150 | 52 | 8 | 57.1 |  |
| **Chronic Disease** |  |  |  |  |  |
| *Yes* | 23 | 8 | 0 | 0 | 0.271 |
| *No* | 265 | 92 | 14 | 100 |  |
| **History of documented prior Covid-19 eposides** |  |  |  |  |  |
| *Yes* | 11 | 3.8 | 14 | 100 | 0.001 |
| *No* | 277 | 96.2 | 0 | 0 |  |
| **Any evidence of pneumonia** |  |  |  |  |  |
| *Yes* | 1 | 0.3 | 3 | 21.4 | 0.001 |
| *No* | 278 | 99.7 | 11 | 78.6 |  |
| **Immunization status** |  |  |  |  |  |
| *Complete* | 265 | 92 | 14 | 100 | 0.546 |
| *Partial* | 21 | 7.3 | 0 | 0 |  |
| *Unimmunized* | 2 | 0.7 | 0 | 0 |  |
| **Family history of documented COVID-19** |  |  |  |  |  |
| *Yes* | 184 | 63.9 | 14 | 100 | 0.005 |
| *No* | 104 | 36.1 | 0 | 0 |  |
| **Number of family members** |  |  |  |  |  |
| *<=3* | 8 | 2.8 | 0 | 0 | 0.583 |
| *4-6* | 173 | 60.1 | 7 | 50 |  |
| *7-9* | 81 | 28.1 | 5 | 35.7 |  |
| *10-12* | 21 | 7.3 | 1 | 7.1 |  |
| *13+* | 5 | 1.7 | 1 | 7.1 |  |

### 3.3 positive long covid-19 patients with most dominant symptoms

(fatigue 12/ 85.7%), (cough 10/ 71.4%), (post exertional malaise 5/ 35.7%) were the most detected symptoms among the 14 positive patients. Followed by (headache, dizziness, hair loss, loss of appetite, loss/change in smell and taste, difficulty in sleep, mood change, abdominal pain, change in bowel habits, chest pain) to lesser extent.

## 4. Discussion

COVID-19, a highly contagious respiratory disease, caused by SARS-CoV-2 virus. The World Health Organization (WHO) established the name “COVID-19,” which stands for “Coronavirus Disease 2019,” in December 2019. In Wuhan, China, has reported the first instances of a novel coronavirus. The virus was eventually called SARS-CoV-2. COVID-19 symptoms can range from mild to severe (fever, cough, fatigue), and can lead to hospitalization and even death, especially in high-risk populations such as the elderly and those with underlying health conditions (23, 24). Shortage of RR PCR for the diagnosis of COVID19 help to spread of COVID19 infection among population (25, 26)

According to current data, the virus transmits through respiratory droplets mostly amongst people who are in close contact with each other, such as at a conversational distance. When an infected person coughs, sneezes, speaks, sings, or breathes, the virus can spread in microscopic liquid particles from their mouth or nose. When infected particles in the air are breathed at close range (this is sometimes referred to as short-range aerosol or short-range airborne transmission) or come into direct contact with the eyes, nose, or mouth, another person might get the virus. (droplet transmission). The virus can also spread in poorly ventilated and/or congested interior environments, where people spend more time. This is due to the fact that aerosols can linger in the air or travel beyond than a conversational distance. (this is often called long-range aerosol or long-range airborne transmission). People can become infected by touching their eyes, nose, or mouth after contacting surfaces or things contaminated with the virus.

There is no single study that had been done in Iraq in which they investing the prevalence of long covid-19 in children, however, in Fars, Iran a cross-sectional study had been conducted to check the prevalence of long covid-19 In children in which they found: out of 58 children 26 of them (44.8%) experienced long covid-19 symptoms. These symptoms included fatigue in 12 (21%), shortness of breath in 7 (12%), exercise intolerance in 7 (12%), weakness in 6 (10%), and walking intolerance in 5 (9%) individuals.

Another study included 267 participants in Italy, showed a percentage lesser than 20% of children were reporting persistence of symptoms for longer duration through an electronic survey. Among these symptoms fatigue was the most frequent.

In comparison to our study these two studies also shows that percentage of long covid-19 in children is still questionable, and variable. However, fatigue seems to be the most frequent symptoms among most of children(27).

Moreover, our study underwent some limitations that should be mentioned: for instance, the children were afraid and crying from needles, which led to disability in checkups and tests., Parents who did not let us do the test for their children, time limitations. In addition to this, our study includes children aged from 5 to 12 years of age. However, a lot of children were aged either above or below this range of age. Together lead to insufficient sample size for statistical measurements.

## 5. Conclusion

The challenges of coronavirus-associated acute respiratory disease called coronavirus disease 19 (COVID-19) are now extending to its long-term sequelae. long COVID-19 is a complex and multifaceted condition that can affect people who have had COVID-19, even if they had a mild or asymptomatic case. Children rarely develop a severe respiratory disease in the acute phase of COVID-19. however, they still were victims to expresses symptoms such as (fatigue, cough, post exposure malaise) after 4 weeks from the active phase of covid-19 had resolved.

4.6% (14 children) among those who were positive for the serological test underwent the challenge of long covid -19 in our study.

## Data Availability

All data produced in the present work are contained in the manuscript

## 6. Recommendation

To address the ongoing challenges of long COVID-19, continued research and collaboration among healthcare providers, researchers, and patients are essential. Long COVID-19 is a public health concern that requires ongoing attention and resources, as well as support for those who are experiencing its debilitating effects. By furthering our understanding of long COVID-19 and developing effective treatments, we can improve the lives of those affected by this condition and mitigate its impact on communities worldwide.

## Notes

### Competing Interest Statement

The authors have declared no competing interest.

### Funding Statement

This study did not receive any funding

